# Cervicovaginal metabolites mediate the relationship between vaginal microbiota and risk for Chlamydia trachomatis infection

**DOI:** 10.64898/2026.11.16.26368223

**Authors:** Smritee Dabee, Mor Tsamir-Rimon, Omolara Akingba, John Kinuthia, James W. MacDonald, Theo K. Bammler, Daniel Raftery, Kevin Hybiske, Ken Tapia, Walter Jaoko, Elhanan Borenstein, Jennifer E Balkus, R. Scott McClelland, Heather B. Jaspan

## Abstract

**Background:** *Chlamydia trachomatis* (CT) remains the most prevalent bacterial sexually transmitted infection and a cause of adverse reproductive outcomes. Epidemiologic studies describe bacterial vaginosis (BV) as a major risk factor for CT infection. Emerging data suggest that BV-associated bacteria and their metabolites contribute to CT susceptibility. Here, we measured the role of specific bacteria in incident CT and evaluated a hypothesized causal role between the vaginal microbiota, its metabolites, and CT risk.

**Methods:** We conducted a nested case-control study of Kenyan women who engage in transactional sex from the Mombasa Cohort who acquired CT versus those who did not, and compared their vaginal microbiome and metabolome at the visit preceding CT acquisition. We determined CT infection and total bacterial load using qPCRs. 16S rRNA sequencing was used to characterize the genital microbiome, and BV status was determined using Nugent-Hillier and Amsel scoring criteria. Metabolites were measured using targeted liquid chromatography-mass spectrometry from a subset of women.

**Findings:** Women with BV, high vaginal microbial diversity, and community state types III and IV were significantly more likely to have CT at their next visit. CT infection itself did not alter vaginal microbial diversity. More specifically, both concentrations and relative abundances of *Lactobacillus crispatus* and several BV-associated bacteria (*Sneathia sanguinegens, Umbribacter vaginalis, Prevotella amnii*, *Hoylesella/Prevotella timonensis,* and *Ureaplasma urealyticum*) were predictive of CT outcomes. Vaginal metabolites differed between cases and controls pre-CT, including Tyramine, reduced and oxidized Glutathione, and sucrose. In a mediation analysis, the vaginal metabolome significantly mediated the relationship between the microbiome and CT infection.

**Interpretation:** We show for the first time that the association between BV and CT risk is mediated by differences in the metabolome. Identification of the mechanisms underlying increased CT risk with BV can inform interventions to reduce infections in high-risk women.

**Funding:** US NIH (R01AI13214401-A1)

**Research in context:** *Evidence before this study:* *Chlamydia trachomatis* (CT) remains the most reported bacterial sexually transmitted infection globally and can lead to serious adverse health outcomes. Previous studies have pointed to an association between the vaginal microbiota composition, particularly the presence of bacterial vaginosis (BV)-associated taxa, and chlamydia susceptibility. However, most studies have been cross-sectional, measuring the microbiome at the time of infection. The few longitudinal studies specifically assessing the microbiome dynamics preceding CT infection have generally been limited by small sample sizes or infrequent sampling. Further, the mechanisms underlying this relationship remain unclear.

*Added value of this study:* In this nested case-control study, we evaluated the vaginal microbiome preceding CT acquisition and compared participants who subsequently acquired infection with matched controls who remained uninfected. We found that women who acquired CT had a significantly higher vaginal microbial diversity in the past 1-2 months, while the CT infection itself did not lead to broad changes in the microbiome, suggesting directionality to this relationship. Participants who acquired CT were >2-fold more likely to have BV by Nugent-Hillier or Amsel criteria at the previous visit. Having a diverse community state type IV was associated with the strongest likelihood of CT acquisition. More specifically, women who acquired CT were more likely to have a microbiome characterized by the depletion of *Lactobacillus crispatus* and higher abundances of *Sneathia sanguinegens, Umbribacter vaginalis, Prevotella amnii*, *Hoylesella (Prevotella) timonensis,* and *Ureaplasma urealyticum.* To assess the possible mechanism through which the microbiome influenced CT risk, we measured the cervicovaginal metabolome in a subset of participants. We found that 70% of the effect of the vaginal microbiome on CT acquisition risk could be explained by the metabolome, highlighting the role of bacterial metabolites in CT susceptibility. Overall, by focusing on samples collected prior to infection acquisition, with a short period between visits, our study contributes robust temporal evidence that the vaginal microbiome impacts CT susceptibility rather than simply being correlated, with the presence of CT, with metabolites potentially playing a key role in this relationship.

*Implications of all the available evidence:* Our findings support the concept that the vaginal microbiome is a potentially modifiable risk factor for CT acquisition, with specific BV-associated bacteria, alongside *L. crispatus* depletion, identified as key modulators of risk. This study is a step towards confirming the directionality of the microbiome-CT infection relationship and establishing potential mechanisms through which the microbiome influences CT risk. These findings, along with previous literature, have the potential to lead to microbiome or metabolite-targeted interventions that would reduce the risk of CT acquisition.

## Introduction

With over 130 million infections globally each year, *Chlamydia trachomatis* (CT) is the most prevalent bacterial sexually transmitted infection (STI)^1^, accounting for approximately 40% of all STIs^1^. Prevalence is highest among adolescent and reproductive-aged women and is linked to serious adverse health outcomes, including pelvic inflammatory disease, infertility, ectopic pregnancy, neonatal infections, and increased risk for HIV acquisition^1^. Preventing CT infection is a global public health priority.

Epidemiologic studies identify bacterial vaginosis (BV), classically characterized by a higher abundance of anaerobic and pathogenic bacteria such as *Gardnerella*, *Fannyhesseae*, and *Prevotella* species, amongst others, as a risk factor for CT infection^2,3^. A growing body of longitudinal and mechanistic studies points to the role of specific BV-associated bacterial taxa in CT acquisition, persistence, and clearance, while *Lactobacillus* species dominance, particularly *L. crispatus,* is protective^4–7^. In young women in the US, a diverse vaginal bacterial community with high relative abundances of *Lachnocurva vaginae* preceded CT acquisition^8^. However, mechanisms driving this relationship are unknown.

BV-associated shifts in the vaginal metabolome may contribute to increased risk of CT. Urogenital CT requires tryptophan for growth and to establish an infection, which it can salvage from indole, a metabolite produced by several BV-associated bacteria^9,10^. More specifically, CT infection correlated with higher vaginal kynurenine/tryptophan ratios and diverse anaerobic microbiota, highlighting an immunometabolic link between microbiome composition and host susceptibility^10^. Loss of *Lactobacillus* dominance corresponds with elevated pH, reduced lactic acid, and increased short-chain fatty acid production, factors associated with mucosal barrier disruption and altered innate immunity^11^. Together, these findings suggest that the cervicovaginal microbiome influences CT pathogenesis by shaping epithelial/immune readiness, nutrient availability, microbial competition, and local metabolite milieu.

To better understand the temporal dynamics between BV and CT, and explore the mechanisms underlying their interactions, we conducted a longitudinal case-control analysis among HIV-seronegative Kenyan women engaged in sex work. We examined the impact of BV, specific bacterial taxa, and their metabolic profiles on the risk of acquiring CT.

## Methods

### Cohort description

Participants in this nested case-control study were enrolled in the Mombasa Cohort, an open cohort in Kenya established in 1993. Cohort characteristics, eligibility criteria, and study protocol have been described previously^12^. Briefly, eligible individuals were of female sex at birth, aged 18–50 years, lived in Mombasa, engaged in transactional sex, and provided informed consent.

This nested case-control sub-study used data collected between September 2010 and November 2021 (suspended from March-July 2020 due to the COVID pandemic) from non-pregnant, HIV-seronegative participants. Although gender identity was not assessed in the Mombasa cohort, it was assumed based on the study protocol that all participants identified as cisgender women. Ethical approval was obtained from the ethical review committees at Kenyatta National Hospital (P197/04/2012) and the University of Washington (STUDY00000615).

### Study procedures and data collection

Structured interviews and physical examinations were conducted at enrollment, collecting demographic, sexual behavior, contraceptive, and sexual history. Participants were followed longitudinally, and during monthly follow-ups, sexual behavior, vaginal washing practices, and symptom data were collected. Pelvic examinations and clinical and laboratory evaluations for STIs were performed. Blood samples were collected monthly for HIV testing.

As part of the larger Mombasa Cohort study, vulvovaginal cotton-tipped swabs (Puritan, USA) were collected monthly for BV evaluation by Nugent and Hillier scoring, and nucleic acid amplification STI testing (NAAT) for CT, *Neisseria gonorrhoea*, and *Trichomonas vaginalis*. Quarterly visit samples were tested for STIs by NAAT, and monthly swabs were tested for BV, vulvovaginal candidiasis, wet mount, and *N. gonorrhea* culture, while the other swabs were stored at -80°C. Syndromic management was used for STI treatment during study visits, with additional treatment provided based on laboratory test results. Additional Dacron dry swabs (Puritan) were stored for multi-omic analyses.

Quarterly visit NAAT results were used to identify incident CT infections. Once identified, stored monthly swabs were tested retrospectively to identify the first CT-positive visit (labeled as Index) and the preceding CT-negative visit (Index-1 visit). These individuals were labeled as cases. Incidence-density sampling was used to randomly identify a set of matching control participants (CT-negative at two consecutive visits) at a 1:2 case: control ratio. Overall, N=77 incident CT cases and N=154 controls were included from 166 unique individuals, of whom 45 contributed more than one distinct case or control sample pair across time (Fig 1).

**Fig 1.**
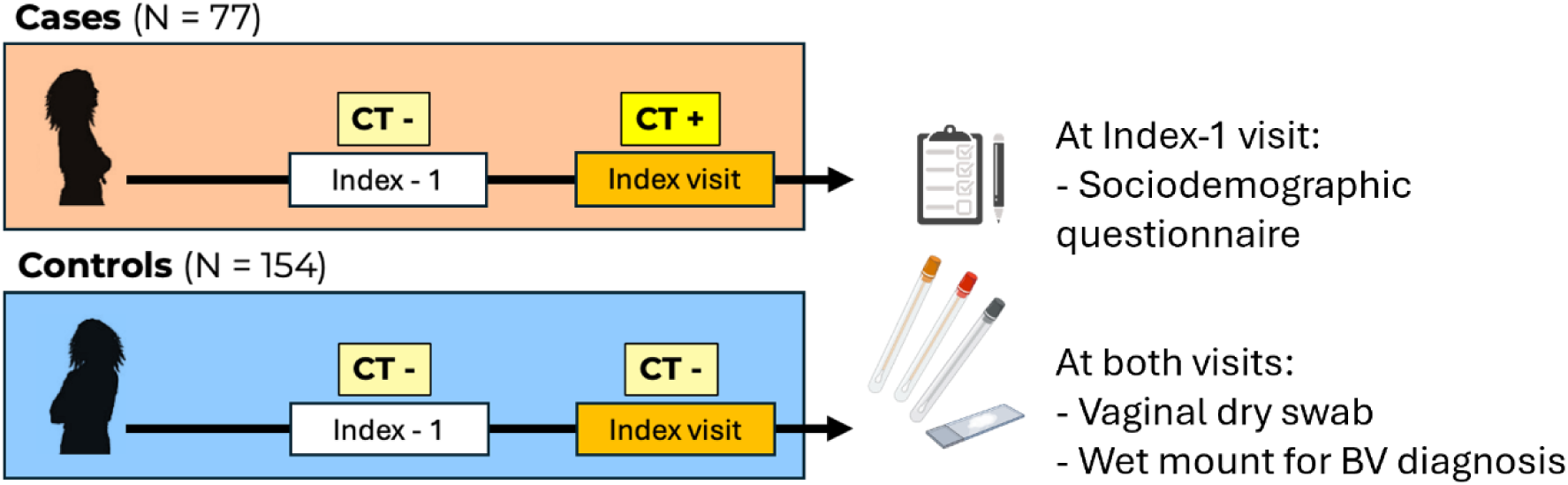
Study design. Sampling strategy for selecting cases and controls in a 1:2 ratio, matched on calendar time using incidence-density sampling. Sociodemographic information was collected at the Index-1, while vaginal swabs and a slide for BV diagnosis by Nugent scoring were collected at both the index and index-1 visits. CT: *C. trachomatis.* Index -1 visit: Visit before *C. trachomatis* acquisition. Index visit: Visit at *C. trachomatis* acquisition.

### Laboratory methods

#### *C. trachomatis* and HIV testing

Cervical specimens were tested for CT using NAAT (Aptima Combo-2, Hologic, USA). HIV-1 testing was performed according to Kenyan guidelines, using Determine (Unipath Limited, UK) and Unigold (Trinity Biotech, USA) kits.

#### BV testing

One vulvovaginal swab was smeared on a slide for a wet mount, which was used to diagnose BV using both the criteria of Nugent-Hillier and Amsel’s criteria^13,14^. Women were categorized as Nugent-Hillier BV-positive (scores 7-10), intermediate (4-6), or negative (0-3). Women were considered Amsel BV-positive if they had three out of four criteria: vaginal pH>4·5, clue cells on wet mount, thin, homogeneous discharge, and a positive whiff test with 10% potassium hydroxide.

#### Bacterial community profiling using high-throughput 16S rRNA sequencing and total 16S qPCR

Stored swabs were thawed, resuspended in sterile PBS, and shaken at 100rpm for 3min. Bacterial DNA was extracted using the Qiagen DNeasy Powersoil HTP 96 kit according to the manufacturer’s instructions. DNA was amplified using primers spanning the V3-V4 region of the 16S rRNA gene as previously described^15^. Amplicons were purified using AMPure XP beads (Beckman Coulter), quantitated using qPCR, and pooled in equal mass amounts. Published mock communities of known abundance (BEI resources), and pure *E. coli* DH10B cultures were included as positive controls to be used in cross-contamination filtering and error rate modeling. Negative controls included both extraction and PCR controls. Paired-end sequencing was performed on an Illumina MiSeq platform using V3 600-cycle kits.

The total bacterial load was determined using the BactQuant assay^16^, a qPCR assay targeting the V3-V4 region, give an estimate of the total 16S rRNA copies per swab.

#### Detection and quantification of the genital metabolome

Broad-based targeted metabolic profiling was performed at the Index-1 visit for a subset of participants who had a second swab available (24 Control and 12 Case participants) using an LC-MS/MS platform (Northwest Metabolomics Research Center). The LC-MS assay targeted 361 metabolic species and 4 stable isotope-labeled internal standards (SILISs).

Aqueous genital metabolites were methanol-extracted using a previously described protein precipitation method^17^. Briefly, 1ml of 80:20 methanol (containing ^13^C_5_-glucose and ^13^C_2_-glutamate)/H_2_O solution was added to frozen dry swabs, vortexed, and stored at -20°C for 30 min. Samples were then sonicated in an ice bath for 10min, vortexed and centrifuged for 15min at 18,000 x g and 4°C. Supernatant was collected from each vial, dried on a Speedvac at 30°C for about 2hrs, and reconstituted in 0·5ml of LC-matching solvent containing 13C2-tyrosine and 13C3-lactate. A 0·3ml aliquot was transferred into LC vials for LC-MS acquisition. Four SILISs were added to the samples to monitor sample prep and instrument performance.

A combination of pooled study samples and commercially available pooled serum was used as quality controls (QCs) to monitor instrument performance and allow batch-to-batch normalization. These QCs were extracted and processed using the same procedure as the samples. Sham swabs without human contact served as negative controls to assess background signals originating from chemical impurities present in the swabs used to collect vaginal fluid. Each QC sample was injected once for every 10 study samples. Measured MS peaks were relatively quantified as area-under-the-curves of the multiple reaction monitoring transitions using AB Sciex MultiQuant 3·0·3 software.

### Data analysis

All amplicon sequence variant (ASVs) calling was performed using DADA2 (v1·32·0) within R (v4·4·0). An updated version of the Silva training set v138 was used for taxonomic annotation, available at https://github.com/SmriteeDabee/Custom-16S-database-for-marker-gene-taxonomic-assignment. Run-specific contamination filtering was performed using microfiltR (https://github.com/itsmisterbrown/microfiltR). Samples with <1000 filtered and annotated reads were discarded.

The phyloseq, vegan, DESeq2, radEmu, corncob, Aldex.2, MaAsLin3, microbiome, tidyverse, epiDisplay, mediation, and gvlma packages were used for ecological comparisons and downstream analyses. Community State Types (CSTs) were assigned using VALENCIA. Differential abundance methods were used to identify taxa found at different fold-changes preceding CT incidence. Wilcoxon Rank Sum tests and permutational ANOVA were used to compare paired diversity measures respectively, and logistic regressions were used to determine odds ratios comparing microbial diversity to CT incidence.

Metabolomics relative quantification values were normalized using a cyclic-loess method using limma. Metabolites with more than 70% missingness were excluded, and the remaining missing observations were imputed using missForest. The estimated imputation error, based on the out-of-bag observations, was 0·26.

Absolute abundances of different bacterial taxa were inferred using the relative abundances generated by 16S rRNA sequencing and the total 16S bacterial burden per swab generated by qPCR. These absolute abundances were used in downstream ordinal regression models. Taxa were agglomerated to their lowest available taxonomic annotation for the differential abundance analyses and regression models. Because microbiome data is compositional, we carried out a centered log ratio (CLR) transformation before generating principal components for the integrative microbiome-metabolome analyses. For the mediation analysis, we selected a bootstrap approach because it does not rely on an assumption of normality and has been shown to be appropriate for smaller samples.^18^

### Data availability

Raw sequence data for 16S rRNA gene amplicon sequences are available under BioProject accession number PRJNA1510256 (pending publication). Additional information can be made available on request.

## Results

The median age was 35 [IQR: 28-44] years, and participants who subsequently acquired CT were younger compared with those who remained CT-negative (median 29 vs 38 years; p<0·0001; Table 1). There were no differences in sexual debut age between cases and controls. The median interval between Index and Index-1 visits was longer for cases (61 days, IQR 32-134) than controls (33 days, IQR 28-42).

**Table 1.** Participant characteristics*.

|  | Controls | Cases |
|---|---|---|
| N | 154 | 77 |
| Age | 38 [31-46] | 29 [25-36] |
| Age at sexual debut | 16 [15-18] | 17 [15-18] |
| Vaginal sex ever | 151 (98%) | 77 (100%) |
| Vaginal sex during menses ever | 22 (14·3%) | 8 (10·4%) |
| Any oral sex | 17 (11%) | 23 (29·9%) |
| Anal sex ever | 2 (1·30%) | 4 (5·19%) |
| Method of contraception at cohort enrollment |  |  |
| None | 22 (14·3%) | 8 (10·4%) |
| Condoms only | 78 (50·6%) | 32 (41·6%) |
| Oral Contraceptive Pill | 4 (2·60%) | 2 (2·60%) |
| Depot medroxyprogesterone acetate (DMPA) | 19 (12·3%) | 15 (19·5%) |
| Intrauterine device (IUD) | 3 (1·95%) | 2 (2·60%) |
| Levonorgestrel implant | 21 (13·6%) | 18 (23·4%) |
| Tubal ligation | 2 (1·30%) | - |
| Hysterectomy | 5 (3·25%) | - |
| BV categories (Nugent and Hillier score) |  |  |
| Negative (0-3) | 96 (62·3%) | 37 (48%) |
| Intermediate (4-6) | 17 (11%) | 6 (7·79%) |
| Positive (7-10) | 41 (26·6%) | 34 (44·2%) |
| BV by Amsel’s criteria | 14 (9·09%) | 16 (20·8%) |
| Vaginal insertion practices in the past |  |  |
| Product used |  |  |
| None | 9 (5·84%) | 6 (7·79%) |
| Water | 60 (39·0%) | 32 (41·6%) |
| Soap | 78 (50·6%) | 32 (41·6%) |
| Other | 7 (4·55%) | 7 (9·09%) |
| Method | 17 (11%) | 6 (7·79%) |
| None | 6 (3·90%) | 6 (7·79%) |
| Fingers only | 112 (72·7%) | 45 (58·4%) |
| Other | 36 (23·4%) | 26 (33·8%) |

\*Numerical variables are reported as Median [Interquartile range], and categorical variables are reported as Frequency (Percentage).

### Women with higher microbial diversity were more likely to acquire *C. trachomatis*

Women with CT at the Index visit had significantly greater odds of having been Nugent-Hillier BV positive at the Index -1 visit compared to controls (BV-neg as reference: odds ratio [OR]=2·15 [95% confidence interval: 1·19-3·90]; Fig. 2). This association was not found among those with an intermediate Nugent-Hillier score at Index-1 (OR=0·92 [0·31-2·40]), although the number of BV-intermediate women was small. Similarly, women with incident CT by the Index visit had significantly higher odds of having Amsel BV at the Index-1 visit (OR=2·62 [1·20-5·78]). Using 16S rRNA gene data, Shannon and Chao1 scores, measures of alpha diversity, were generated as metrics of microbial diversity. CT acquisition was associated with about two-fold higher odds of having higher α-diversity at the previous visit (OR=1·98 [1·14-3·49] and 2·21 [2·27-3·90], respectively).

**Fig 2.**
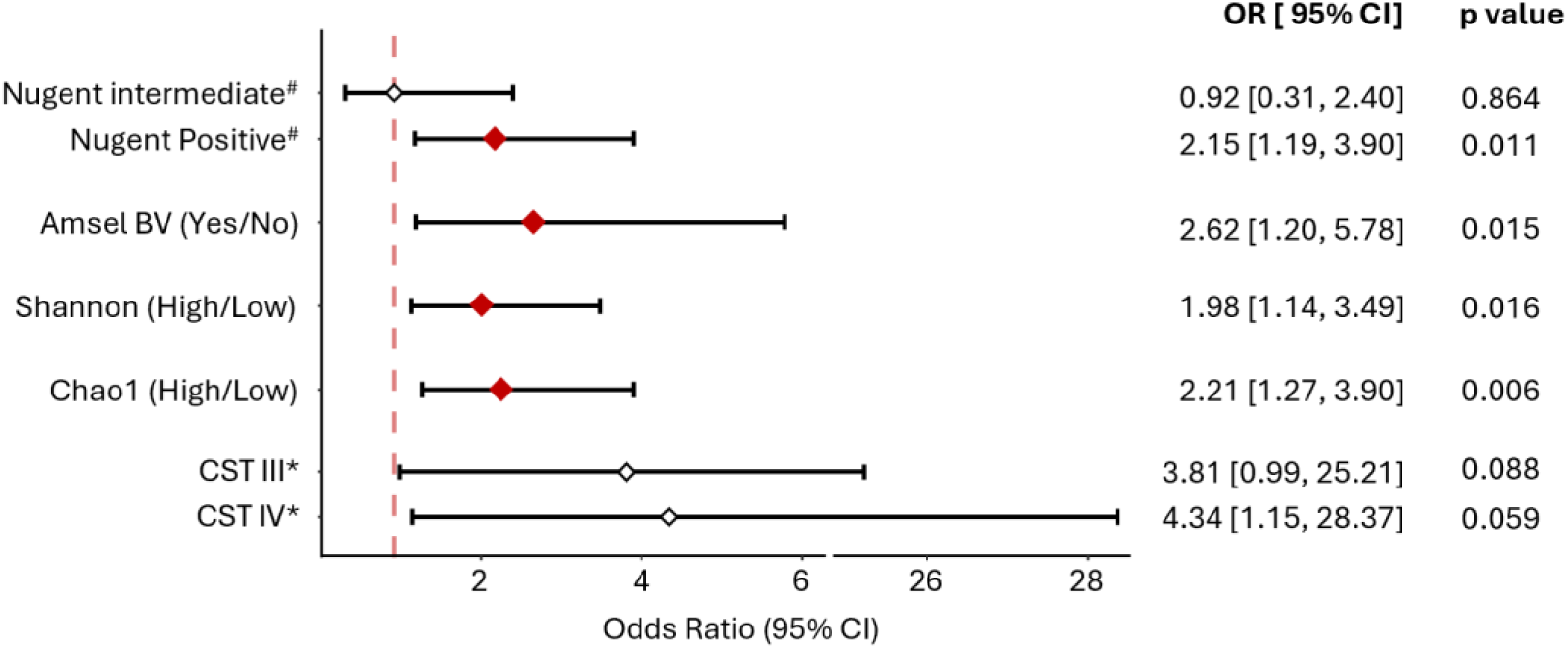
The overall genital microbial composition and odds of *C. trachomatis* acquisition risk at the next visit. OR: Odds ratio; BV: Bacterial vaginosis; CST: Community state type

The genital microbiota of women was categorized into CSTs (Fig S1). The odds of having a CST-IV microbiome was 4·34-fold higher in women who acquired CT when compared to individuals with an *L. crispatus*-dominant microbiota (CST-I) (95% CI:1·15-28·37; Fig 2). The odds of having *L. iners*-dominance (CST-III) were also 3·81-fold higher with CT acquisition (95% CI: 0·99-25·21). Within CST-IV subgroups, those who acquired CT were more likely to have a microbiota dominated by *Gardnerella vaginalis* and *Fannyhessea vaginae* (CST-IV-B vs CST-I; OR: 4·41 [0·95-20·35]), although this association was not statistically significant.

**Fig S1.**
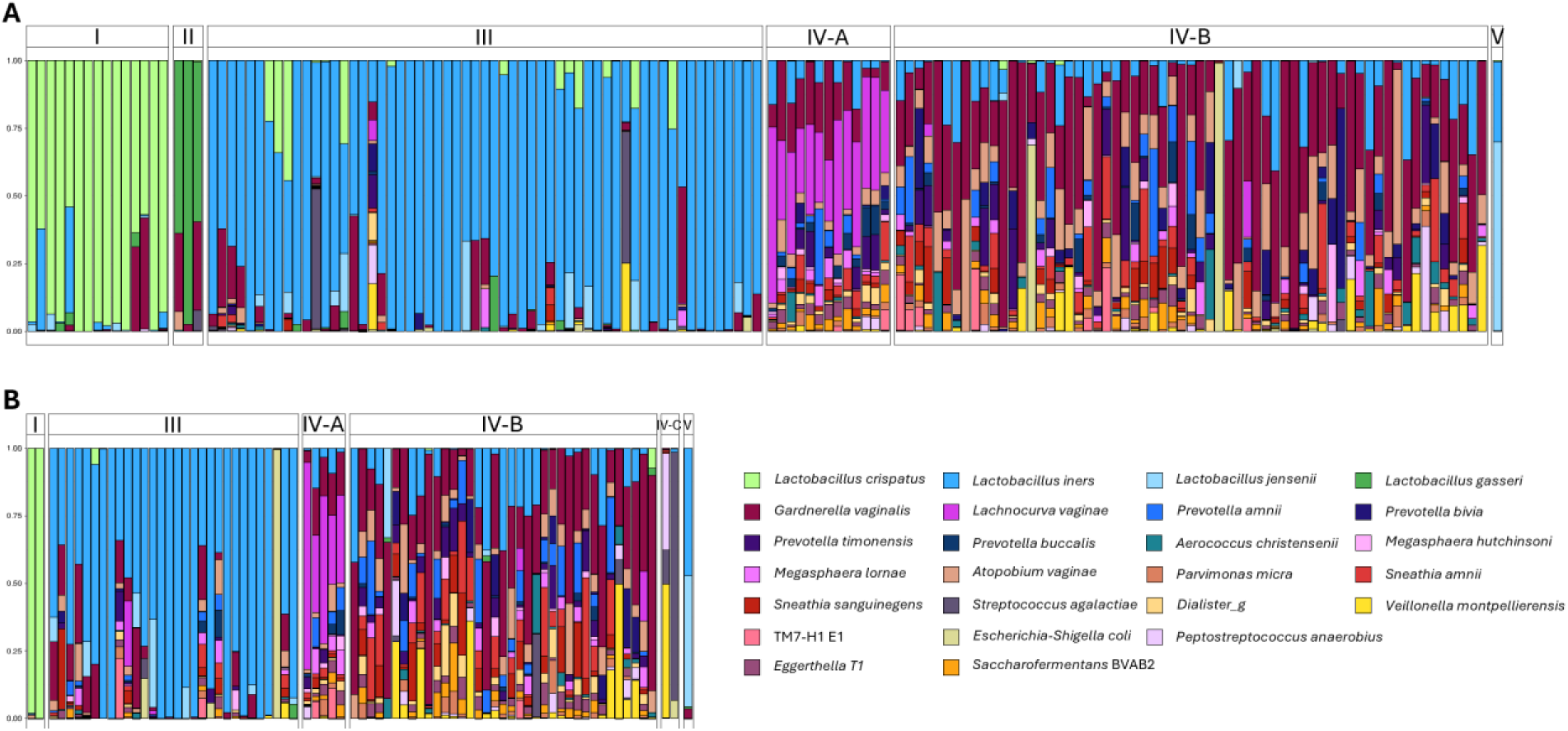
Microbial composition categorized by community state types (CST) at the Index-1 visit for (**A**) women who remained negative for *C. trachomatis*, and for (**B**) those who acquired *C. trachomatis* at the subsequent Index visit

### Acquiring *C. trachomatis* did not change the overall vaginal microbiota

To begin establishing the directionality of the relationship between vaginal dysbiosis and CT, we examined differences in microbial diversity between cases and controls, and between visits. As in Fig 2, at the pre-CT visit (Index-1), we found that the microbiota was significantly more diverse among women who subsequently acquired CT, compared to control participants (Fig 3A; Chao1 diversity: p=0·02; Shannon diversity: p=0·01). This was accompanied by a significant difference in community composition between cases and controls at the Index-1 visit (Bray-Curtis distance: PERMANOVA p=0·03; Fig 3B). However, among CT cases, there was no shift between visits in either within-participant (Chao1: p=0·44; Shannon: p=0·62; Fig 3C) or between-participant (Bray-Curtis distance: p=0·58; Fig 3D) microbial diversities, indicating that vaginal dysbiosis precedes CT infection. The difference in microbiome diversity between cases and controls persisted at the incident visit (Index) after CT acquisition, with a more diverse microbial composition among the cases (Chao1: p<0·01; Shannon: p=0·05) (Fig 3E and F).

**Fig 3.**
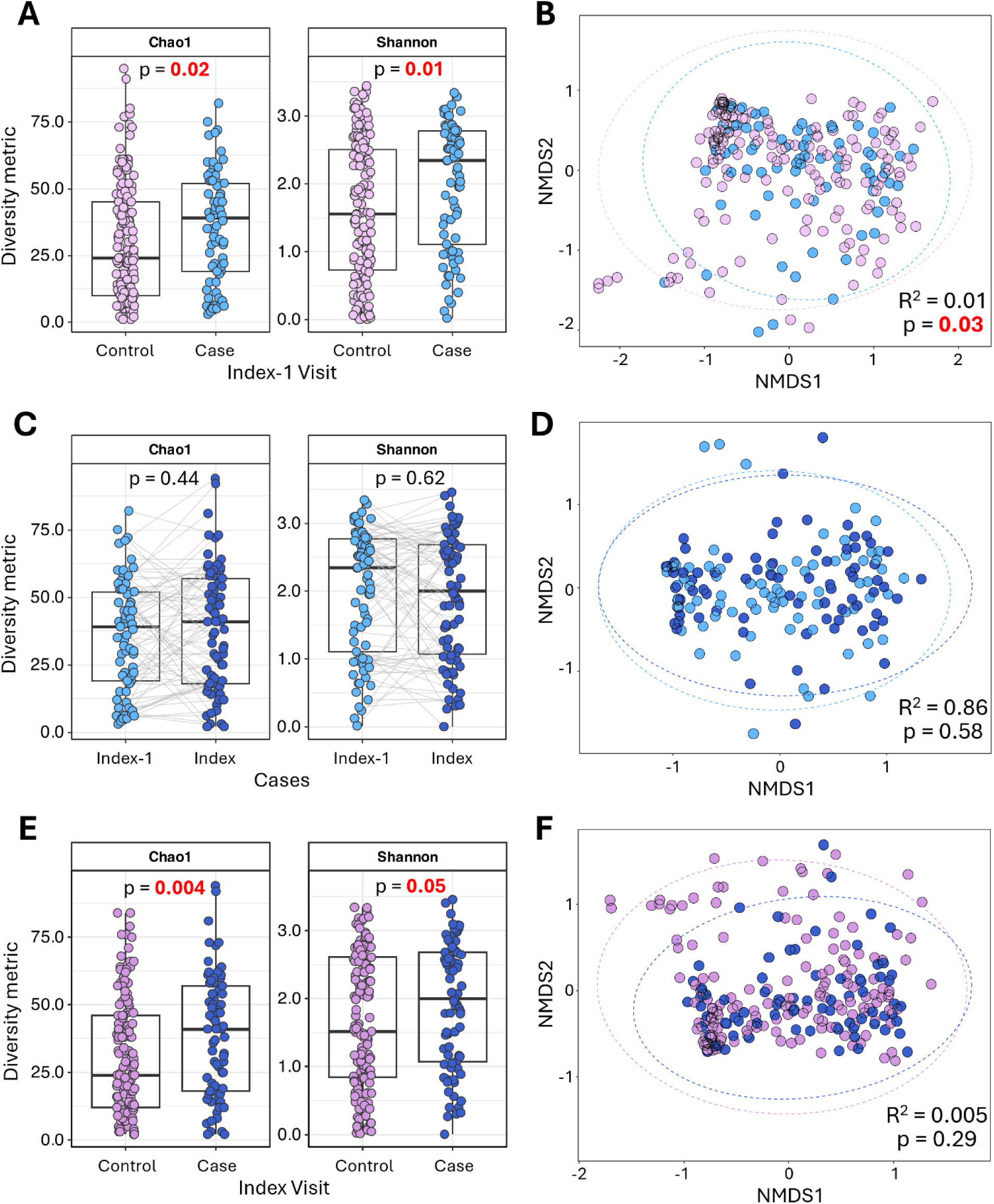
Genital microbiome diversity across visits and participants. Comparing Chao1 and Shannon alpha diversities and Bray-Curtis beta diversity (**A** and **B**) between cases and controls at the visit preceding CT infection, (**C** and **D**) between the preceding visit and the incidence visit among cases, and (**E** and **F**) between cases and controls at the incidence visit. P values ≤005 were considered statistically significant.

### *C. trachomatis* infection is preceded by the absence of *L. crispatus* and high relative abundances of specific BV-associated bacteria

Next, we assessed whether specific bacterial taxa at the Index-1 visit were associated with subsequent CT infection. We compared fold-change (FC) differences in normalized abundances of individual taxa between cases and controls. Given the high variability in methods available to calculate differential abundance, with most methods having some shortfalls, we used multiple methods: DESeq2, Aldex.2, MaAsLin3, RadEmu, and corncob. Only taxa significantly differentially abundant with ≥3/5 methods, after false discovery rate correction within each method, were considered meaningful (Fig 4). Women who subsequently acquired a CT infection had an average of 2·05 log-FC lower *L. crispatus* (4/5 methods: DESeq FC = -4·13; Aldex.2 FC: = -1·26, MaAsLin3 FC = - 1·64; RadEmu FC = -1·20) and >0·88 - 1·46 log2 FC increases in seven BV-associated taxa: *Sneathia sanguinegens, Umbribacter vaginalis, Prevotella amnii*, *Hoylesella (Prevotella) timonensis,* and *Ureaplasma urealyticum* (Fig 4). Other BV-associated taxa found at higher FC among cases were *Dialister microaerophilus* (corncob, Aldex.2), *Parvimonas micra* (Aldex.2), and *Granulicatella elegans* (MaAsLin3).

**Fig 4.**
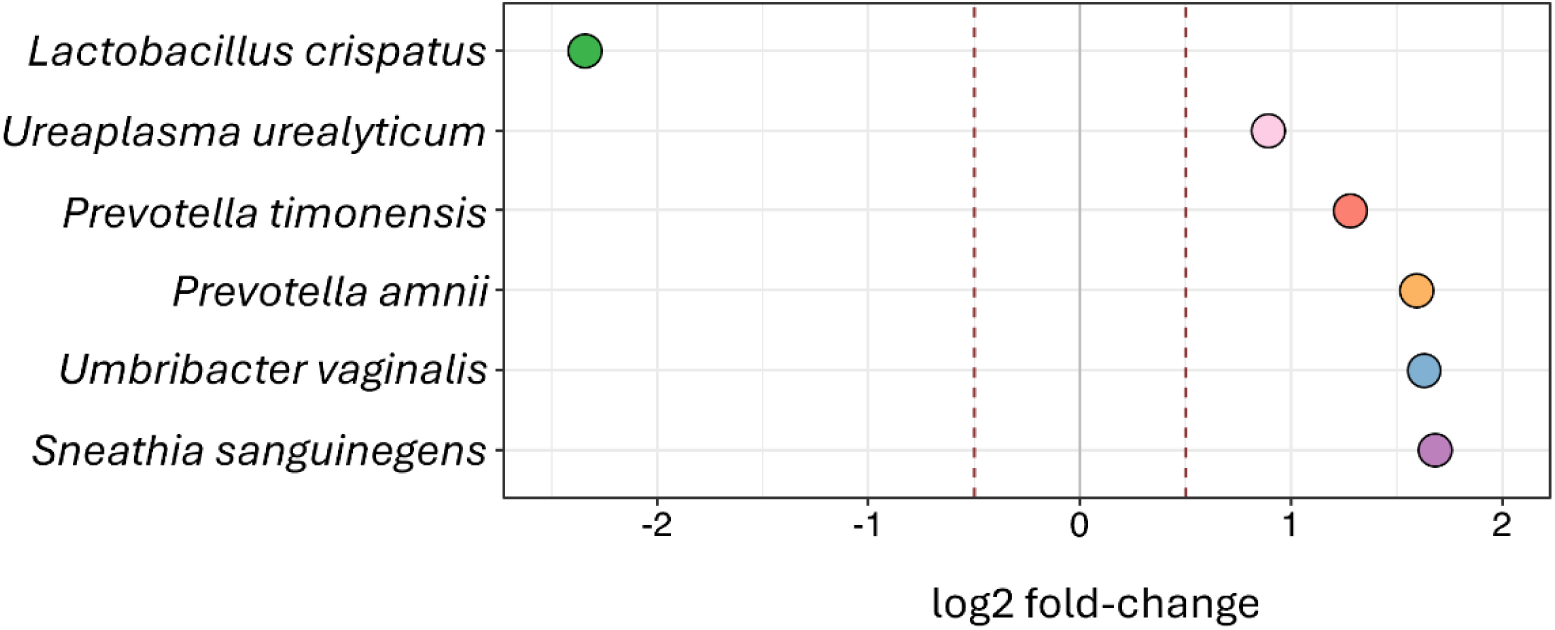
Fold-change differences in the abundance of specific bacterial taxa among those who subsequently acquired *C. trachomatis* compared to those who remained *C. trachomatis*-negative. Taxa that were differentially abundant using 3/5 methods (DESeq2, Aldex.2, MaAsLin3, RadEmu, and corncob). The red dotted lines represent a 0.5-fold change in abundance. Only differentially abundant taxa with a p ≤ 0.01 after adjusting for multiple comparisons were included.

We assessed the impact of the absolute concentration of these identified taxa on a woman’s overall odds of CT acquisition. The absolute inferred bacterial concentrations were categorized as Undetectable/ Tertile 1/ Tertile 2/ Tertile 3. The odds of CT acquisition in the presence of each differentially abundant taxon were estimated using ordinal logistic regressions. A subsequent CT infection was associated with a 53% decrease in the odds of being in a higher *L. crispatus* tertile at Index-1 (OR = 0·47 [0·23 - 0·89]; p = 0·03; Table 2), while CT incidence was positively associated with being in a higher tertile for the other identified BV-associated bacteria, with between a 1·72- to 2·08 higher odds (all p<0·05; Table 2), highlighting the protective role of *L. crispatus* and that specific BV-associated taxa may be implicated in risk, possibly due to their specific metabolic contributions to the vaginal environment.

**Table 2.** Ordinal logistic regressions showing the association of the differentially abundant taxa with *C. trachomatis* cases versus controls.

| **Taxa** | **Tertiles** | **Log 10(absolute abundance) (median [IQR])** | **Positive for CT at next visit (N= 230) [n (%)]** | **OR [95% CI]** | **p value** |
|---|---|---|---|---|---|
| *Lactobacillus crispatus* | Undetectable | 0 [0-0] | 63 (27·4%) | Ref |  |
|  | Tertile 1 | 3·56 [3·11-3·94] | 7 (3·04%) | 0·47 [0·23-0·89] | 0·03 |
|  | Tertile 2 | 5·18 [4·7-5·68] | 5 (2·17%) |  |  |
|  | Tertile 3 | 6·78 [6·52-7·10] | 2 (0·87%) |  |  |
| *Prevotella amnii* | Undetectable | 0 [0-0] | 39 (17·0%) | Ref |  |
|  | Tertile 1 | 4·18 [3·43-4·70] | 12 (5·22%) | 2·08 [1·22-3·57] | 0·01 |
|  | Tertile 2 | 5·63 [5·38-6·10] | 15 (6·52%) |  |  |
|  | Tertile 3 | 7·07 [6·76-7·27] | 11 (4·78%) |  |  |
| *Hoylesella (Prevotella) timonensis* | Undetectable | 0 [0-0] | 30 (13·0%) | Ref |  |
|  | Tertile 1 | 3·88 [3·47-4·50] | 14 (6·09%) | 1·91 [1·15-3·17] | 0·01 |
|  | Tertile 2 | 5·18 [4·93-5·47] | 19 (8·26%) |  |  |
|  | Tertile 3 | 6·26 [6·10-6·92] | 14 (6·09%) |  |  |
| *Sneathia sanguinegens* | Undetectable | 0 [0-0] | 33 (14·3%) | Ref |  |
|  | Tertile 1 | 3·92 [3·44-4·45] | 13 (5·65%) | 1·90 [1·13-3·17] | 0·01 |
|  | Tertile 2 | 5·51 [5·33-5·66] | 20 (8·7%) |  |  |
|  | Tertile 3 | 6·65 [6·30-7·12] | 11 (4·78%) |  |  |
| *Ureaplasma urealyticum* | Undetectable | 0 [0-0] | 35 (15·2%) | Ref |  |
|  | Tertile 1 | 3·16 [2·75-3·52] | 16 (6·96%) | 1·89 [1·12-3·17] | 0·02 |
|  | Tertile 2 | 4·15 [3·99-4·32] | 15 (6·52%) |  |  |
|  | Tertile 3 | 5·02 [4·79-5·48] | 11 (4·78%) |  |  |
| *Umbribacter vaginalis (Eggerthella* T1) | Undetectable | 0 [0-0] | 29 (12·6%) | Ref |  |
|  | Tertile 1 | 3·93 [3·36-4·51] | 14 (6·09%) | 1·72 [1·04-2·85] | 0·03 |
|  | Tertile 2 | 5·21 [5·07-5·48] | 20 (8·7%) |  |  |
|  | Tertile 3 | 6·38 [6·22-6·90] | 14 (6·09%) |  |  |

### Metabolites associated with CT acquisition

The baseline metabolome (Index-1 visit) was measured in a subset of participants (n=36). While there was no statistically significant clustering of the vaginal metabolome between cases and controls at the Index-1 visit (Fig 5A, Partial Least Squares Discriminant Analysis p>0·05), the metabolites with the highest VIP scores that contributed the most to principal components (PC) 1 and 2 loadings (i.e. explaining the most to variability in clustering) included the proteins/amino acid-related compounds: Tyramine, N-Ac-Glutamate, N-Acetyl-Aspartate (NAA), 5-Aminovaleric Acid, Histamine, and Sarcosine; thiols: Reduced Glutathione (GSH) and Oxidized glutathione (GSSG); sugars: Sucrose, Glucose, Glucose 6-phosphate (G6P), and UDP-Glucose; the amine Cadaverine; nucleotide/nucleoside: Guanosine monophosphate (GMP), Inosine, Guanosine, and Uridine; lipids: Glycerophosphocholine, isoValerylcarnitine, and Deoxycarnitine; the vitamin Thiamine, and the co-enzyme Nicotinamide adenine dinucleotide (NAD) (Fig 5B).

**Fig 5.**
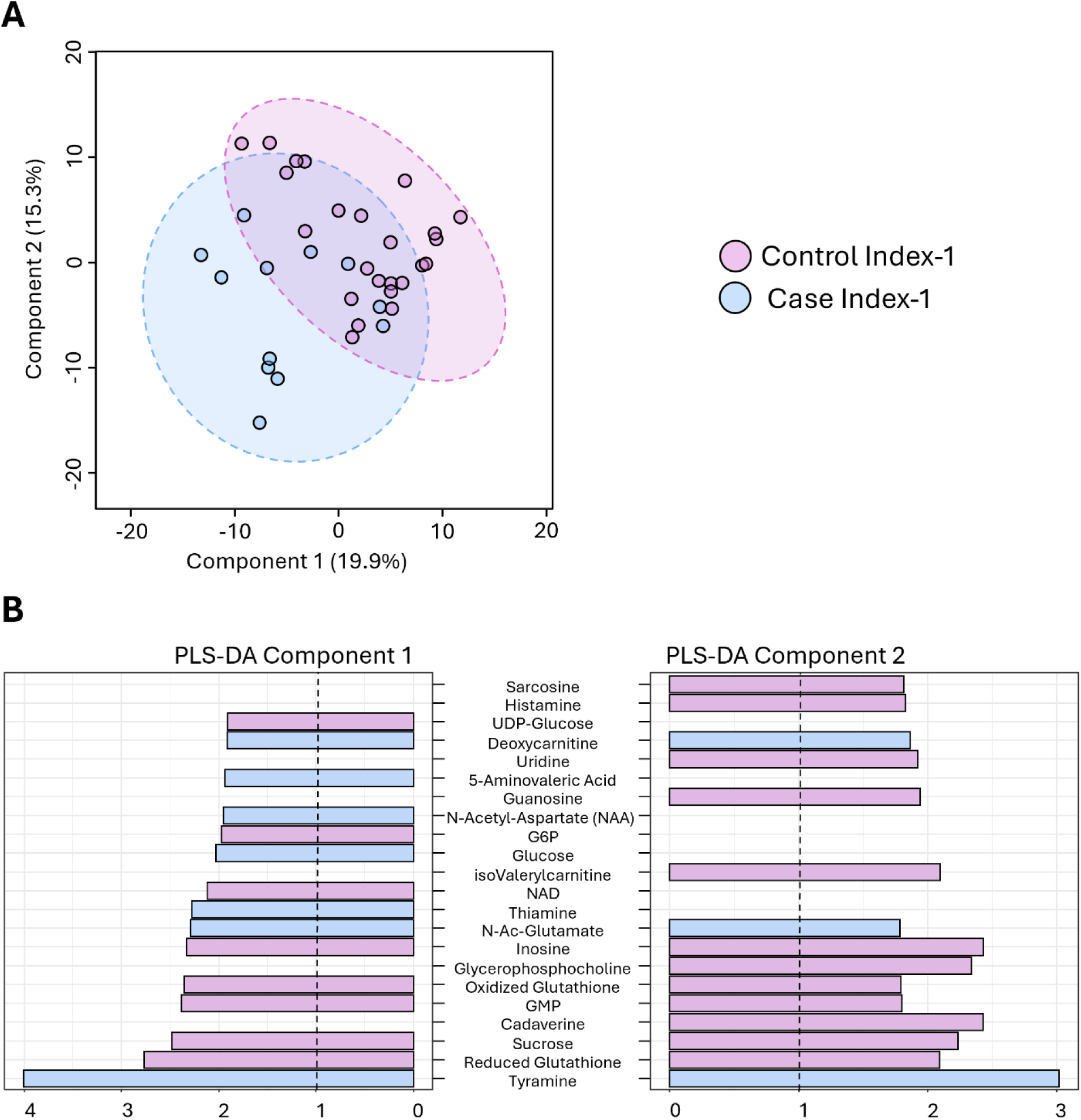
(**A**) Partial Least-Squares Discriminant Analysis (PLS-DA) showing the clustering by metabolome between cases and controls at the Index-1 visit. The x- and y-axes indicate the contribution of component scores 1 and 2 to the overall variability (**B**) Bar plot of Variable Importance in Projection (VIP) scores for the top 15 metabolites with the largest contributions to component scores 1 and 2, respectively. A VIP score >1 was considered significant for identifying discriminatory metabolites.

### Mediation analysis using the overall microbial composition and all metabolites

To assess whether the vaginal microbes were altering risk for CT via their metabolites, we performed a causal mediation analysis (Fig 6). The loadings on PC1 from the PLS-DA (Fig 5B) were used along with PC1 from a PLS-DA carried out using CLR-transformed microbial features (Fig S2). The total effect was decomposed into the average causal mediation effect (ACME; indirect effect through the metabolome) and the average direct effect (ADE, effect not operating through the metabolome). We found a strong mediation effect of the microbiome on the genital metabolome, which, in turn, was statistically significantly associated with a woman’s CT acquisition risk (Average causal mediation effect [ACME]: *β*=-0·03 [-0·05--0·01], p=0·02; proportion mediated: 70·4%, p=0·03), indicating that the increased risk of CT acquisition with altered vaginal microbiota was substantially mediated by the vaginal metabolome.

**Fig 6.**
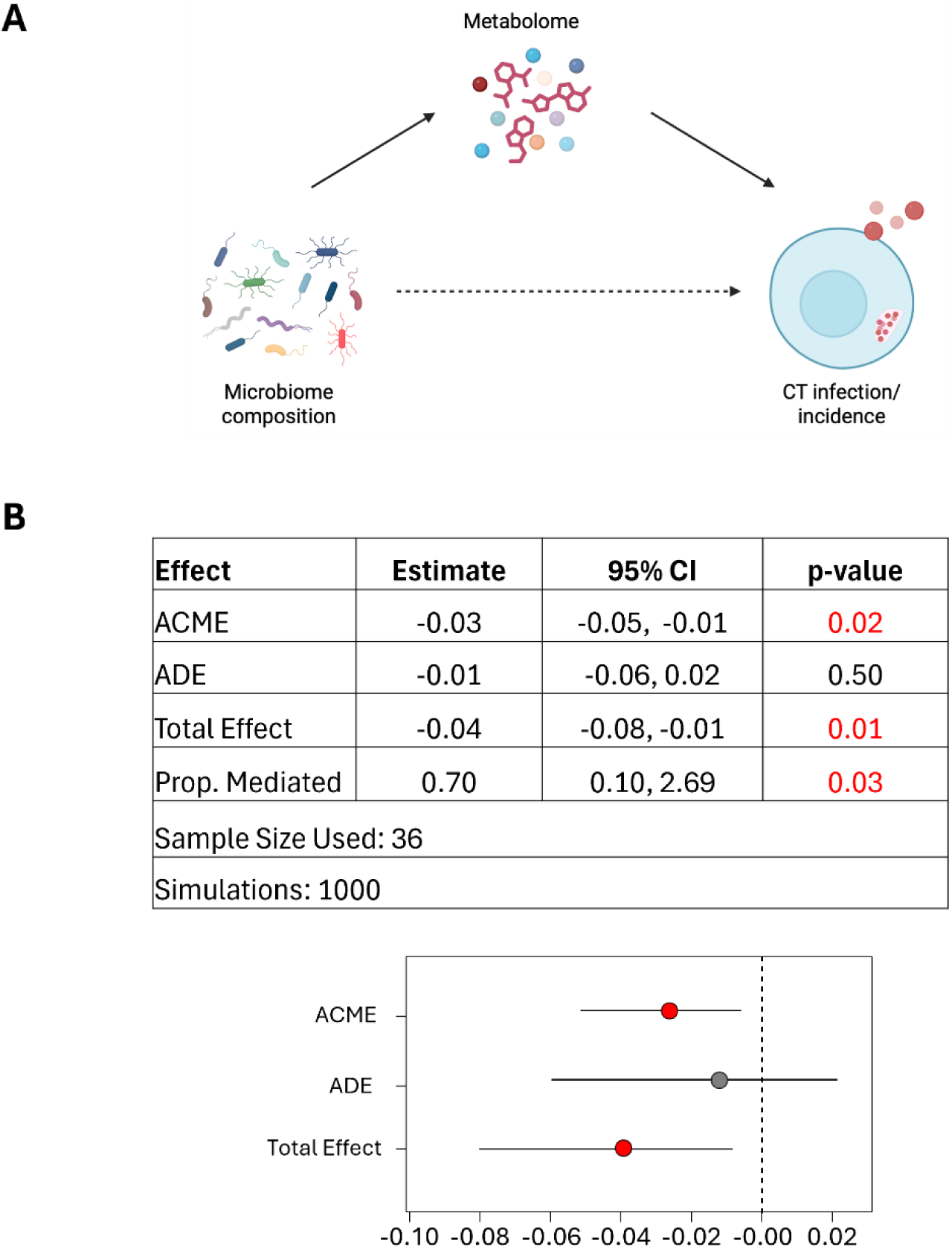
(**A**) Hypothesized model of Ct acquisition where the microbiome composition influences Ct risk through the metabolome (**B**) Mediation analysis evaluating the extent to which the genital metabolome (PLS-DA PC1) mediates the relationship between the CLR-transformed microbiome composition (PLS-DA PC1) and Ct incidence. The total effect was decomposed into the average causal mediation effect (ACME; indirect effect through the metabolome) and the average direct effect (ADE, effect not operating through the metabolome). The proportion mediated is the ratio of the ACME to the total effect. Point estimates and 95% confidence intervals are shown, after nonparametric bootstrapping, and p values <0.05 are considered statistically significant.

**Fig S2.**
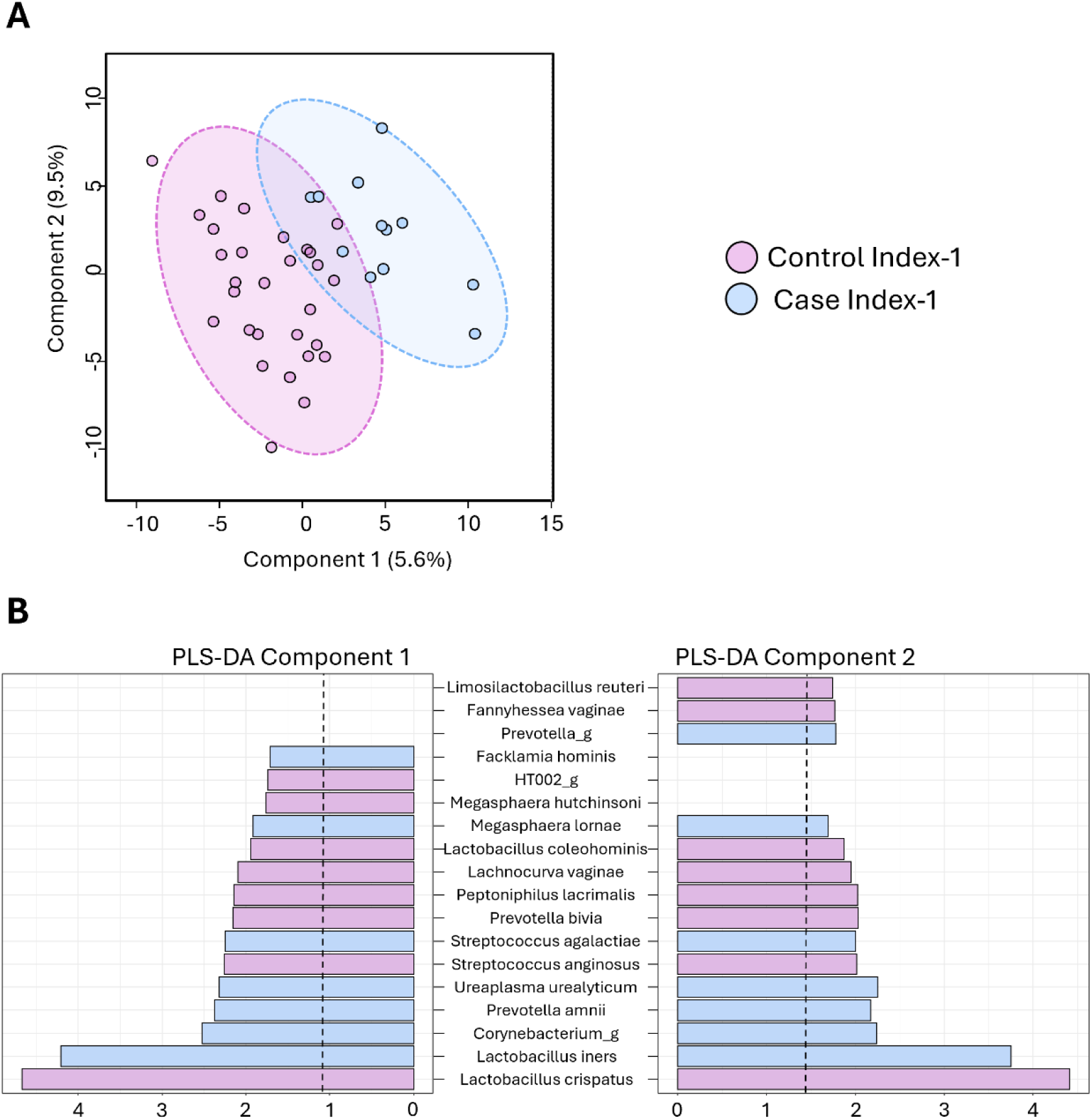
(**A**) Partial Least-Squares Discriminant Analysis (PLS-DA) showing the clustering between cases and controls, based on their microbiome composition, at the Index-1 visit in the subset of women with matching metabolome data (N = 36). The x- and y-axes indicate the contribution of component scores 1 and 2 to the overall variability (**B**) Bar plot of Variable Importance in Projection (VIP) scores for the top 15 bacterial taxa with the largest contributions to component scores 1 and 2, respectively. A VIP score >1 was considered significant for identifying discriminatory metabolites.

## Discussion

Multiple past studies have pointed to a link between CT infection and BV, but the causal relationship remained unclear, primarily as both are influenced by sexual behavior. It is increasingly clear that behavioral factors are insufficient to explain the increased risk of CT acquisition and that biological factors are key. In this cohort of Kenyan women at high risk for STIs, with frequent follow-up visits^12^, we show that acquiring a new CT infection was associated with significantly higher odds of having high vaginal microbial diversity in the past 1-2 months. The concentrations of specific taxa were strongly associated with subsequent CT acquisition. Further, we demonstrate that CT acquisition does not change the overall microbiome composition, indicating a directional relationship where the vaginal microbiome at the time of CT exposure may be an important determinant of infection. Finally, we found that the effect of the microbiota on CT acquisition may be mediated via its metabolome.

Two prior studies have examined the vaginal microbiome prior to incident CT, although these studies had longer periods of time between the prior and the incident visits (6-12 months) than in this current study. In a cohort of young Dutch women, having an *L. iners*-dominant microbiota was associated with a 2·6-fold higher odds of acquiring CT within the year^19^, while a CST-IVA microbiota, but not CST-IVB or CST-III, was associated with a 2·51-fold increased odds of CT incidence among adolescents and young women six months before infection^8^. Here, by measuring the odds of subsequent infection within 2 months, we can substantially advance our understanding of the direction of this relationship. We found a higher prevalence of BV, both by Nugent-Hiller scoring and Amsel criteria (with a >2-fold increase in odds), among women who subsequently acquired CT. Interestingly, this association was not present among those with an intermediate Nugent score. Along these lines, women were more likely to acquire CT if they had higher within-participant microbial diversity at the previous visit. These increased odds appeared to be particularly associated with having a CST-IV microbial composition, with a >4-fold increase in CT incidence. Furthermore, while women who acquired CT had a more diverse microbiome at baseline, the CT infection itself did not have an impact on the overall diversity, providing evidence that microbial diversity preceded infection.

The difference in microbial diversity between those who acquired CT compared to those who remained negative was driven by higher relative abundances of specific BV-associated bacteria, but to a larger degree, by the depletion in *L. crispatus*, where CT infection was preceded by a >2-fold lower relative abundance of *L. crispatus*. The increased odds of CT acquisition were highest with increases in the abundances of *P. amnii, H. timonensis* and *S. sanguinegens* respectively, while the protective effect of *L. crispatus* was associated with 53% lower odds of infection. The increases in these BV-associated taxa are like those described by Usyk et al (2025), who also report higher differential abundances of *Prevotella* and *Sneathia* spp., amongst others approximately 6 months before infection. They did not report differences in *Lactobacillus* spp, although this may be due to the dynamic nature of the vaginal microbiota over time. The fact that *Prevotella* spp. are strongly associated with risk raises the possibility that metabolites preferentially produced by this genus could enhance CT infectivity. For example, *Prevotella* spp. have high sialidase activity^20^, which may facilitate CT infection.

In line with this, CT acquisition was associated with the presence of several metabolites at the pre-infection visit. Among those, GSH, GSSG, sucrose, glucose, G6P, GMP, guanosine, uridine, and NAD have previously been described as being associated with CT. GSH may allow CT to remain infectious, while GSSG was involved in persistent infection^21,22^. CT can leverage the host metabolism to access sugars needed for its growth and survival – it can induce increased glucose transporter expression (GLUT1/GLUT3) and, in HeLa cell cultures, higher sucrose concentrations led to increased stability of chlamydial membranes, longer viability of infectious elementary bodies (EBs), and a higher number of chlamydial inclusions^23,24^. G6P is a preferential source of energy for EBs, and in culture, GMP contributes to CT growth^25^. Due to its reduced genome, CT relies on its host for its biosynthetic pathways and requires substrates like cytidine, uridine, and guanosine for survival^26^. Other metabolites, particularly cadaverine, deoxycarnitine, and 5-aminovaleric-acid, could also be a consequence of the concurrent presence of bacterial dysbiosis and inflammation, but not necessarily directly associated with the CT infection itself^11,27,28^. Interestingly, contrary to our hypothesis that indole or tryptophan would be permissive to CT infection due to its reliance on these host-derived substrates^9,10^, we found no association between these metabolites and subsequent CT infection.

The vaginal microbiome was highly predictive of the overall metabolome composition. Importantly, 70% of the effect of the microbiome on CT risk was explained by the cervicovaginal metabolome, tying bacterial metabolites as the mechanism through which woman’s risk of CT infection increases with BV. While other studies have shown that the genital microbiome and the metabolome are highly correlated^29,30^, and while the mediating role of the metabolome on the microbiome has previously been alluded to in previous literature^11^, our study adds considerably by suggesting an influence of the microbial metabolome on CT susceptibility.

A key strength of this study was its case-control study design, with short intervals between visits. However, the small number of women with overlapping microbiome and metabolome data was an important limitation. Nevertheless, our findings suggest that we show that the cervicovaginal microbial dysbiosis may play a substantive role in CT acquisition risk within the subsequent two months with taxon-specific influences and potentially mediated by microbial metabolites. These findings may lead to interventions that could decrease risk of CT infection in women.

## Data Availability

The sequencing reads generated during this study have been deposited in the NCBI Sequence Read Archive (SRA) under BioProject accession number: PRJNA1510256. Additional de-identified information can be made available on request.

## Author contributions

Conceptualisation: JEB, RSM, HBJ, EB; Supervision of data collection: JK, WJ; Data curation: SD, OA, JWM, KT; Formal analysis: SD, MT, JWM; Funding acquisition: JEB, RSM, HJB; Methodology: JEB, RSM, HBJ, EB; Resources: JK, DR, WJ; Supervision: JEB, RSM, HBJ, EB; Writing – original draft: SD, HJB; Writing – review C editing: OA, DR, KH, EB, JEB, RSM

All authors reviewed the manuscript and agreed with the submission of this manuscript.

## Acknowledgements

This study was funded by the US National Institutes of Health (R01AI13214401-A1).

